# Staff Perspectives on Implementing Opt-Out Blood-Borne Virus Testing in English Emergency Departments: A Qualitative Study

**DOI:** 10.1101/2025.08.01.25332816

**Authors:** Siobhán Allison, Tom May, Jonathan Roberts, Rachel Hill-Tout, Stephen Hindle, Matthew Hickman, Lucy Yardley, Rachel Todd, Robyn Heath, Jeremy Horwood

## Abstract

**Background:** A significant challenge to achieving global 2030 elimination goals for Blood-Borne Viruses (BBVs) is identifying undiagnosed individuals and relinking those who are no longer in care. To address this, the UK government has implemented opt-out BBV testing in Emergency Departments (EDs) to increase access to BBV testing in high prevalence areas. All adult ED patients having a routine blood test are automatically tested for HIV, hepatitis B and C, unless they opt-out. This study aimed to identify barriers and facilitators to the implementation of ED opt-out BBV testing and provide recommendations for future rollouts.

**Method:** Semi-structured interviews with 23 staff members across five ED sites in very high HIV prevalence areas were analysed thematically, informed by Normalisation Process Theory.

**Results:** While there was some variation in staff knowledge and understanding of the programme, overall acceptance of the opt-out testing approach was found to be high. Training had a positive impact on staff understanding of the purpose of the intervention and the correct process, including the opt-out model. High workloads and competing priorities in EDs were significant barriers to testing. However, some specific systems and processes that facilitated the uptake of testing included automation and BBV champions. Giving the programme time to embed into practice and ensuring feedback loops and flexibility to ‘tweak’ the process was also essential to sustaining the programme.

**Conclusion:** To embed opt-out testing into emergency care, sites should implement automated test ordering, staff training, clear communication, and dedicated champions, which can help to support earlier diagnosis, reduce inequalities and improve patient outcomes.

## Introduction

The UK government has set a goal to reach both national and World Health Organisation (WHO) targets of zero new HIV transmissions and eliminating Hepatitis C (HCV) and Hepatitis B (HBV) as a public health threat by 2030. A key challenge in reaching these targets is identifying individuals who remain undiagnosed. An estimated 4,700 people in England are living with undiagnosed HIV [1] and 76,820 people remain undiagnosed with hepatitis B virus (HBV) [2]. In 2024, approximately 40% of HIV diagnoses in England were made at an advanced stage of HIV disease (CD4 count below 350 cells/mm³)[3], which increases the risk of hospitalisation and mortality, reduces life expectancy, and results in higher treatment costs [4, 5]. Prompt HIV diagnosis and initiation of antiretroviral therapy reduces viral load, improves life expectancy [6], and prevents onwards transmission [7]. Both HBV and HCV can lead to chronic liver disease and cirrhosis and antiviral treatment can cure most cases of HCV [8].

New and innovative approaches are needed to reach undiagnosed populations. Emergency Department (ED) opt-out blood-borne viruses (BBV) testing (hereafter, “the programme”) is one such approach. Opt-out BBV testing involves all adult ED patients who are having a blood test as part of their care automatically receiving an additional blood test for BBVs, unless they opt-out. Patients are informed via posters, leaflets, and video screens about the testing.

EDs are well-positioned to implement opt-out BBV testing due to their high patient volumes and their ability to reach people underserved by traditional health services [9]. These populations often face significant health inequalities, making robust systems for linkage to care essential [10]. The opt-out testing model also has the potential to reduce the incidence of missed opportunities of diagnosing BBVs [11].

The UK government allocated funding for ED opt-out testing in two waves; Wave 1 in April 2022 to 33 sites in areas with very high HIV prevalence in England (defined as >5 per 1000 in the local population) (across London, Manchester, Blackpool, and Brighton) and Wave 2 in April 2024 to 45 sites in areas with high HIV prevalence across England (defined as >2 per 1000 in the local population).

Evidence from previous ED BBV testing implementation pilots have identified barriers to testing, which are attributed to staff not offering the test rather than patients actively opting out, in fact, there is high public acceptability of BBV testing [12, 13]. Traditionally, BBV testing focused on patient risk factors (including country of birth, sexual behaviour, and history of injecting drug use). Evidence suggests that healthcare workers’ assumptions or unconscious biases may, in some cases, affect whether BBV testing is offered to patients in a routine and equitable way [14, 15].

HIV care has traditionally been provided separately from other medical services, with enhanced privacy and confidentiality [16]. This approach emerged in response to significant stigma faced by people living with HIV, a phenomenon often referred to as the’exceptionalism’ of HIV [17]. As a result, many general NHS staff have had limited experience treating individuals living with HIV or accessing current information about care and treatment, which can contribute to outdated and stigmatising views [16]. In contrast, universal opt-out testing removes the need for such judgments, helping to normalise BBV testing and reduce stigma [15]. The introduction of changes in practices needs to ensure that staff are well supported and aware of the rationale behind the opt-out approach [12].

There is overlap between populations at risk of HIV and HBV and HCV supporting the public health opportunity and importance of combining the three BBV tests in this programme. Early pilots of opt-out testing in EDs focused solely on EDs testing for HIV, missing the opportunity to diagnose viral hepatitis. For example, one study showed that, a single week of testing in an ED revealed that 54 viral hepatitis diagnoses would have been missed if testing had been limited to HIV alone [18].

ED presents unique challenges related to testing implementation, uptake, and connecting patients to ongoing care, compared to other BBV screening programmes in antenatal and prisons. The current study aimed to evaluate the implementation by investigating ED staff views and experiences of opt-out ED BBV testing, as well as the facilitators and barriers to implementation, and provide recommendations to sites.

## Methods

### Study design

The research employed a qualitative design using semi-structured remote interviews with 23 members of staff involved in the delivery of opt-out BBV testing in Wave 1 sites. Interviews were conducted between April and September 2024.

### Participants and recruitment

Staff were recruited from five ED sites in areas with very high HIV prevalence. Sites were selected based on their geographical location, organisational practices, and opt-out testing models/processes. Recruitment involved site Principal Investigators (PIs) informing eligible staff members about the project, either by email or face-to-face, and placing recruitment materials in staff areas (e.g., staff rooms). Staff were eligible to participate if they were (1) employed by a site situated in a very high HIV prevalence area that were implementing and delivering the ED opt-out testing programme, and (2) a member of staff involved in the implementation and delivery of ED BBV testing, from blood collection to laboratory testing to management. We were particularly interested in speaking to patient-facing staff who were taking the blood, such as healthcare assistants and nurses. The recruitment materials included a QR code linking to a web page with study information and a section for interested staff to log their contact details. A member of the research team later contacted them to arrange a suitable interview time. Purposive sampling was used by the researchers to select a diverse range of staff based on their role (e.g., ED staff, laboratory staff, care pathway facilitators) and level of experience (including time in post and current NHS band).

Analysis was conducted alongside recruitment, and the data was continuously assessed to ensure that the study objectives were being met and ‘information power’ had been achieved [19].

### Data Collection

Interviews were conducted by SA and TM via telephone or video call. Written consent was obtained through an online (Qualtrics) consent form prior to the interview, which the participant completed. At the start of each interview, the researcher confirmed that the consent form had been completed and that the participants had no further questions. A flexible topic guide was developed, informed by Normalisation Process Theory (NPT) [20, 21] and included open-ended questions to elicit views on opt-out testing, including its acceptability, perceived organisational barriers, aspects that worked well and areas for improvement. All interviewees were offered a £10 voucher as a thank you.

### Theoretical Framework

NPT is an implementation theory used to understand how new interventions are adopted, embedded, and sustained in routine healthcare practices. NPT proposes that the implementation of interventions is dependent on the ability of participants to fulfil four criteria presented in Figure 1. NPT informed data collection and analysis by guiding exploration of how staff made sense of, engaged with, operationalised and appraised the programme [20, 21].

**Figure 1:**
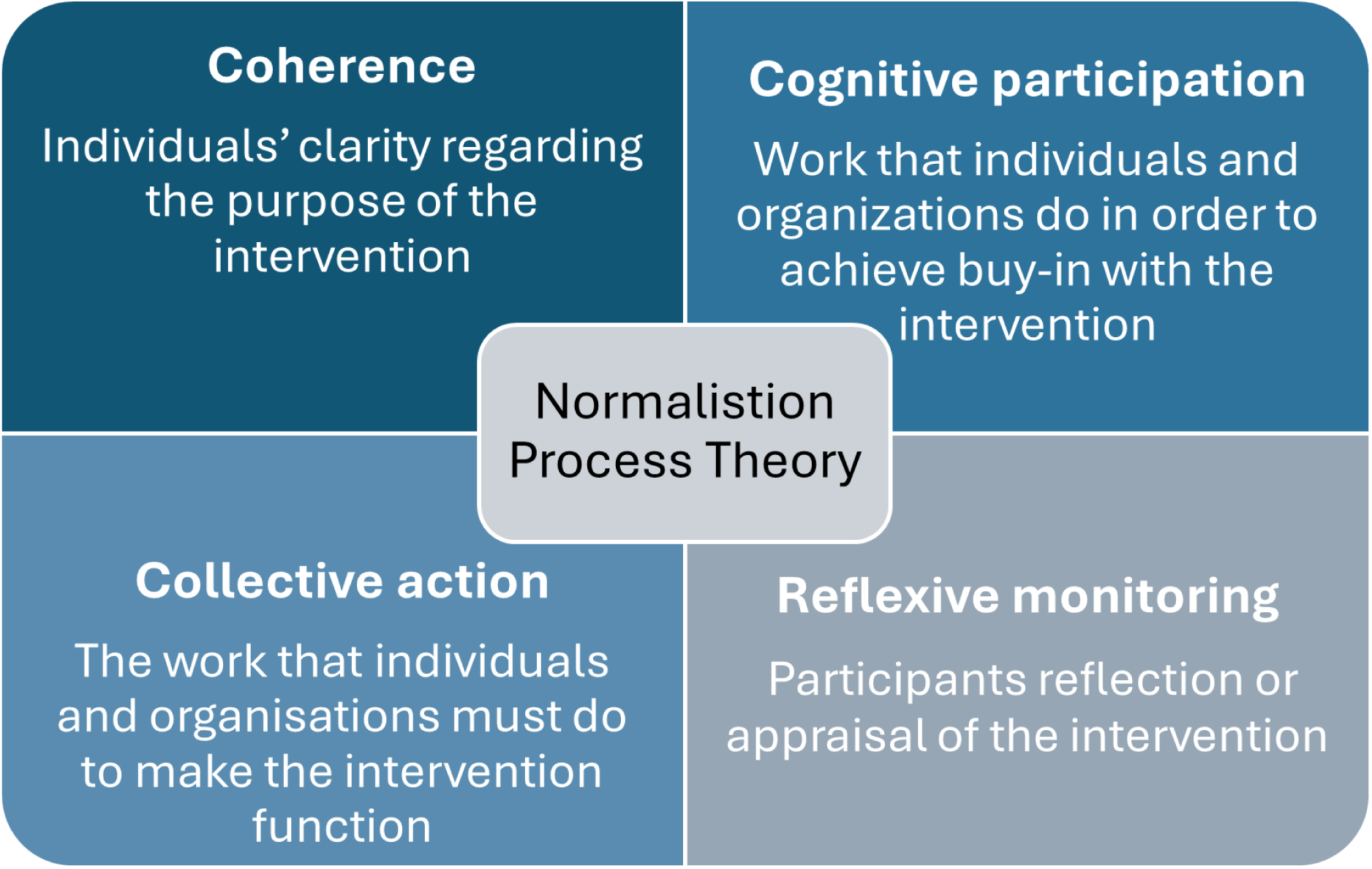
Outlines the NPT constructs [21]

**Figure 2:**
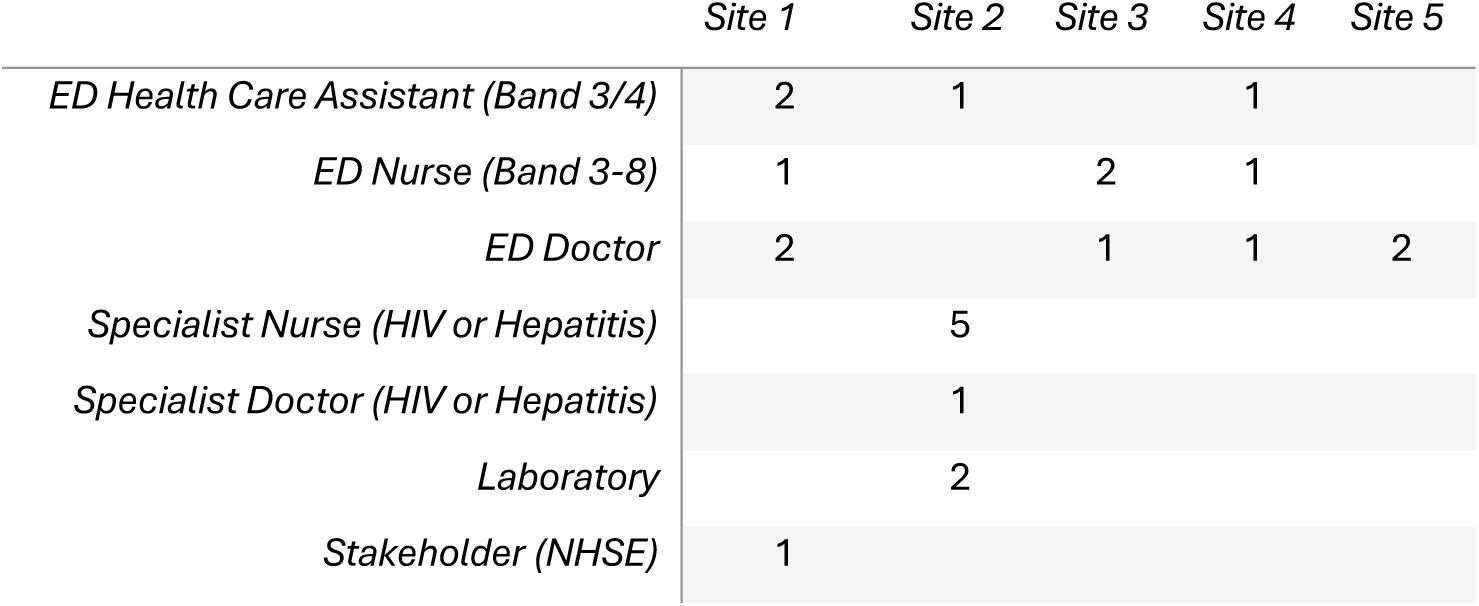
Outlines the roles of the staff who were interviewed

### Data Analysis

With participant consent, interviews were audio recorded, transcribed, anonymised, and uploaded to NVivo V.12 for analysis. Thematic analysis, using a data-driven inductive approach [22], was applied to identify and examine patterns and themes significant to participants and across the dataset. Three transcripts were initially inductively coded independently by two researchers (TM and SA) who and discussed any new codes of potential significance to the research objective. TM and SA are experienced qualitative health researchers, educated to at least a postgraduate level.

TM is a research fellow with a background in social and behavioural science, and SA is a senior research assistant who is also an HIV nurse but has no connection to any of the sites or individuals involved in this study. Analysis began with open coding to generate an initial coding framework. This was then refined through NPT constructs to further investigate developing themes across the data set. The analysis was conducted concurrently with the interviews and was iterative, allowing for further development of the topic guide and coding framework. The coding framework was then applied to the remaining transcripts by SA. Codes were built into broader categories through comparison across transcripts and higher-level recurring themes were developed informed by the NPT constructs. The multidisciplinary research team discussed the data and development of themes to ensure credibility and confirmability, with relevance to the research objectives.

### Findings

Twenty-three interviews were conducted with staff from five sites involved in delivering the programme, including one with a senior NHSE stakeholder overseeing the project. Interviews lasted between 14 and 45 minutes (average 30 minutes). Staff represented a range of roles related to the delivery of the programme.

Themes developed from the analysis aligned with four primary NPT constructs, which are used to structure the findings below, alongside verbatim quotes.

## Coherence

### Purpose of the intervention

Many staff understood the purpose of the intervention, the programme’s benefits to patients and contributions toward public health elimination goals:

> *I think it’s really good. Especially from the point of view of hepatitis C, a lot of people have been found and diagnosed through ED that would otherwise potentially never have been tested…because they’re not necessarily within a demographic that would be targeted for testing, so I think it’s been really positive (Hep C Nurse, Site 2)*

However, some individuals misunderstood the programme’s purpose. For example, one healthcare assistant described the reassurance that opt-out testing provided in the context of potential needle stick injuries:

> *For somebody who’s continuously in contact with patients and handling sharps, it is comforting to know that if I accidentally end up with a needle stick injury, most of my patients agree to have the test, and then I know that I’m…if I’m going to be okay or if there’s any risks (ED HCA, Site 1)*

### New concepts

Most staff appeared to support the introduction of the programme. However, not all fully understood the concept of opt-out testing, including that patients were informed via posters and verbal consent was not required. Patient-facing staff were therefore required to familiarise themselves with new concepts associated with an opt-out model:

> *And there were a couple of people [ED staff] that said not sure if patient consents to it [testing]…which I suppose there’s still a question from staff about what is the consent process? I can’t remember exactly how it was phrased but it was that they weren’t quite sure if proper consent had been done (ED Doctor, Site 1)*

Some staff believed that patients should be informed that they were having a BBV test, and that verbal consent should be sought. This was often due to a lack of familiarity with the opt-out model, including the shift away from requiring verbal consent:

> *I’m not doing [BBV test] without consent– you know – because I dunno what’s gonna happen. Is my name on that? (HCA, Site 1)*

For others, normal practice involved obtaining verbal consent for all blood tests, including BBVs:

> *Oh of course, we always gain consent when – we gain consent for even before getting bloods, we always gain consent if they’re happy for us to take the bloods (ED Nurse, Site 3)*

The need to change previously learned behaviours regarding which regular blood tests they would be requesting posed a barrier to the programme’s delivery, particularly among specific staff groups:

> *And that is a really hard thing to do if people, you know, it’s changing someone’s learnt behaviour that they always just request their normal bloods and go and the sample. Even though it’s such a simple extra it’s hard to change that little bit of behaviour, especially in doctors I feel (ED Doctor, Site 1)*

### Value in the intervention

Staff recognised the potential value, benefits, and importance of implementing the programme. This was often aligned with existing evidence of its effectiveness or benefit to patients:

> *We’ve been told like there are – like we have had people who have been tested, and they’ve come back positive, and it’s changed their lives in the positive, in the fact that they’ve got, in some cases, early diagnosis (HCA, Site 1)*

For ED staff in particular, it was seen as an opportunity to contribute to population-level preventive medicine, an area they felt was often overlooked in emergency care:

> *It’s something I don’t think A&E does enough of, is preventative medicine. (ED Doctor, Site 4)*

### Cognitive participation

#### Key individuals

Implementation demanded considerable time and effort to set up from the service level down to the ward level, including IT and laboratory services. Key individuals played a central role in ensuring the successful implementation of BBV testing. Some sites had formal roles such as champions. For others, it was carried out as an informal role in addition to their existing workload. Both were key in supporting implementation efforts:

> *One of the HIV clinical nurse specialists has really taken a lead (HCV Nurse, Site 2)*

These key individuals helped to foster participation across different staff groups:

> *We’ve developed, kind of ED opt-out champions in the emergency department. So we’ve got a handful of healthcare assistants and nurses who were really almost, kind of empowering to role model the whole service and really encourage it…I think, working more with the BBV champions over there will just make the process more streamlined (HCV Nurse, Site 2)*

#### Buy in

Education and training were key components in ensuring staff engagement and buy-in, as understanding the purpose of the intervention helped staff to engage with it:

> *But once the kind of process and theory behind it was explained, I think that they [staff members] were very much on board(ED Doctor, Site 1)*

Staff discussed how the intervention and training had shifted their perceptions of BBVs. By implementing an opt-out method, the focus shifted away from individual risk onto the importance of universal testing:

> *I can remember sort of older people in resus having heart attacks, and you take their blood and it…you’d find out later that they’re HIV positive and it wasn’t the ones you’d expect, you know…the young homosexuals or the drug users…those people stand out as maybe being HIV positive. So I think it’s very useful to think that…anybody could be HIV positive (ED Consultant, Site 4)*

### Collective action

#### Performing intervention tasks

The intervention required staff to order, collect, and send an additional blood sample to the laboratory. In some sites, BBV tests were automatically included in the standard blood set, the electronic ordering system (EOS) included the BBV test in all blood orders. However, staff remained responsible for physically collecting the sample. Initial training was valuable in ensuring staff understood the importance of including the test. To maintain uptake particularly at sites without automated ordering where staff had to manually request the BBV test via the EOS staff huddles, post-it notes, and other verbal and written reminders proved effective:

> *It’s also being done [reminder about testing] by word of mouth…and then they continuously updated in our handover in the early morning before we start our shift (ED Nurse, Site 5)*

It is recommended that patients who return to the ED within a 12-month period do not undergo a repeat BBV test unless there is a clinical reason. Ideally, the ordering systems would automatically ‘block’ the test being ordered. If blocking is not in place, patients who attend the ED regularly may have unnecessary tests, incurring extra cost and time. Not all sites had this mechanism in place, and staff raised this as a concern:

> *And that would also skew your results, because all the tests that you do, like a chunk of them will be in the same patient within a certain time frame…I don’t think there’s any sort of blocking in A&E (ED Doctor, Site 3)*

Some ED staff highlighted potential positive adjustments to their daily practice, including the removal of the need to risk-assess patients, along with the potential biases associated, which helped to simplify their workflow:

> *I’m not having to double-think it and say ooh, is this the type of person that might have it? Cause, for one, I know that there is no specific type of person, and two, I know that, yeah, that’s not expected of me, just send it (ED Doctor, Site 1)*

### Confidence in the Opt-Out Model

The opt-out model relied on the accessibility of the posters explaining the intervention to patients. Some staff had confidence in the posters and relied on them to inform patients:

> *We just let the posters do the talking (ED Nurse, Site 1)*

In contrast, other staff were concerned that patients had not seen or understood these posters, and therefore sought verbal consent from patients for each individual test:

> *They [patients] could have not seen the poster and they don’t know they’re being tested for HIV, so it could be that we should mention it when we say, ‘We’re taking blood. We’re also testing for HIV (ED Doctor, Site 3)*

Good working relationships and links between the different departments involved (HIV, Hepatitis, and laboratories) ensured the programme continued to run smoothly and allowed issues to be resolved quickly:

> *But I think overall, it works well. I think there’s good communication between hepatology, sexual health, the labs. We often sort of meet and chat, and sort of like tweak things to see how we can get them to work better. So, I think that yeah, it is working well (HCV Nurse, Site 2)*

Collaboration across specialist teams also provided support and advice during the initial stages, which was useful for responding to any modifications to the programme (e.g., integration of HCV testing):

> *When it came to adding in hep C testing…I introduced our HIV and our hep C leads for testing, so that they could work together, because they had so many commonalities. And, actually, that’s been brilliant because er it just means that they can work synergistically and save an awful of time (HIV Doctor, Site 2)*

### Consistent messaging and training

There were inconsistencies both between and within sites regarding whether training was provided and who received it. This led to staff not having the confidence to speak to patients about follow-up care they may receive if they had a reactive test:

> *None of us actually know what the next steps are for that patient. So we kind of say to them…that’s something if you bring that up to your doctor, your doctor will be able to give you more advice on. But us personally don’t know the next steps to be able to advise the patients on what the next steps would be if they was to be tested positive (HCA, Site 1)*

Staff were, at times, only given limited information from senior staff about what was required of them to carry out testing:

> *You were told in passing [about BBV testing] that you know, to see whether they had anything and it was just a screening to catch people that didn’t know (ED Doctor, Site 5)*

### Workforce and context

EDs experienced high volumes of patients, and staff experienced high workloads, which impacted delivery of BBV testing. Sites reported high staff turnover alongside shift work, with some staff only working night shifts. This led to some staff lacking accurate information about opt-out testing:

> *It’s not going well with doctors because we only see new doctors. When we have locums doctors…they don’t know. So I think the doctor needs to have more knowledge about this. Because every week someone came to me, what is red for? Oh it’s for HIV. Oh, so you have to do it for everyone? Yes you have to do it for everyone? You don’t have to ask just do it…so I’m sure most of them they don’t really…know about this (HCA, Site 2)*

There were some initial concerns about the additional workload associated with introducing another blood test:

> *Initially, it kind of felt a bit like that [extra work] because it was another label, another blood bottle…But now it’s just like I said, automatic (Nurse, Site 4)*

### Reflexive monitoring

#### Feedback and impact stories

When sites provided staff with feedback on number of BBV tests conducted, new diagnoses, and personal stories, staff found these messages engaging and felt motivated by the positive impact of the tests:

> *I think it’s the patient stories that were more convincing, rather than the numbers, to sort of say here’s someone actually who came here, you know, actually a new diagnosis who was started on treatment that never, you know, wouldn’t have known without this project, I think that’s quite, quite a powerful teaching tool (ED Doctor, Site 1)*

#### Staff reflections

Staff reflected on the effectiveness of the programme and offered recommendations to enhance future rollouts. Key suggestions included comprehensive training, ensuring staff are well-informed about follow-up care and able to address patients concerns and providing regular feedback including patient stories:

> *Yeah, so the training, have somebody, you know, to go through the training with you. But also have somebody who’s living with the virus, do a talk and meet some of the staff (HCA, Site 4)*

Others suggested having a champion role, collaborating with experienced sites, fostering teamwork and engagement:

> *I think it is probably good to have a dedicated member of staff for that role, because sometimes […] it can be time intensive. So, I think to have that established person with that role, being aware of what their role is within that is an important thing. (HCV Nurse 2, Site 2)*

Overall, staff encouraged new sites to integrate the opt-out testing approach into routine practice:

> *Just do it. You know you’re gonna get reluctance by staff. You’re gonna get reluctance by patients so just do it. Just do it and just get it over with and it’ll become be a routine, like routine so we just crack on. It’s automatically done so yes. (ED nurse, Site 1)*

Staff reflected that there was potential to achieve more through the programme. Stigma and a lack of knowledge amongst patients were highlighted as an issue. Staff considered whether more could be done in educating patients and increasing understanding of BBVs. One participant suggested providing patients with information about BBVs:

> *I think if we come with some, leaflets, to give to them when they’re leaving, after the blood test…they can read about HIV, why it’s so important having bloods, and then why is the treatment. So I think if we give them to the patient at the end…they will be more knowledgeable about what we’re doing (HCA, Site 2)*

However, this did not seem feasible to some staff. Even though they recognised the benefits, they questioned whether ED was the right environment:

> *I don’t know whether A&E is the best place to give education. You know, we have lots of patient leaflets that we hand out…but you walk outside the hospital and the bin’s full of them (ED Consultant, Site 4)*

Others suggested other forms of health promotion ideas, such as videos on tv screens in the ED and other forms of communication, to increase awareness of BBVs and the programme:

> *TV running with some up to date information and maybe some links attached to that (ED Consultant, Site 4)*

### Personal sense of accomplishment and purpose

Overall, being part of the programme for some individuals was important and gave them a sense of accomplishment:

> *It’s a sense of achievement because that’s what we’re trying to do, we’re trying to reach elimination (HCV Nurse, Site 2)*

### Initiating change

ED staff took ownership of identifying issues and making changes to improve the intervention. For example, an HCA identified a problem with staff forgetting to fill the second yellow-top vial and described modifications their site had introduced to address this:

> *With the two yellow you used to get confused before. People, especially the doctors, they used to send one and they thought it’s including HIV in the other bloods. So…I said, can we change, can we do something here so we can void, so they can see one different colour. Which the red I think it’s more…yes it’s red, it’s more obvious (HCA, site 1)*

## Discussion

This study builds upon existing evidence on the acceptability and effectiveness of ED opt-out testing by identifying qualitative insights into the implementation of opt-out testing in EDs [10, 12, 15, 18, 24–29]. Findings identified widespread support and acceptance of BBV testing among staff, despite initial concerns regarding its delivery but also some variation in how staff understood and conceptualised opt-out testing. Staff reported how, over time, training and key individuals helped foster staff buy-in, and the programme became routine once fully implemented and established in their EDs.

Collaboration across teams also played a key role in refining the programme and sustaining staff engagement. Regular feedback on results and impact stories promoted staff reflection on the impact of the programme and helped to maintain staff engagement. This study provides further context to the interim data from the Public Health Evaluation of the programme, which indicates promising results for the programme [30] by showing the acceptability for the programme amongst staff and what barriers may need to be addressed to increase test coverage.

Overall, acceptance of the opt-out testing approach was found to be high, which aligns with existing evidence on staff acceptability of opt-out testing in EDs, staff were particularly focused on the value to individuals’ health [15, 29]. Nevertheless, this study identified how the intervention required a change in staff practices as the opt-out model differed from previous blood taking routines. This demonstrates how fostering a strong understanding of the programme among staff through training and regular staff meetings not only enhances adherence to the model but helps to normalise the process of additional BBV testing [29].

The exceptionalism of HIV testing remains a barrier to testing [12, 25, 31]. Some staff in this study still viewed HIV testing as ‘different’ to other blood tests they took, with the view that explicit consent was needed [12, 25]. This ambiguity is echoed in the literature [24], but as the opt-out model becomes more established, staff confidence is expected to grow [26]. Training impacted positively on staff understanding of the purpose of the intervention and the correct processes, including the way patients were informed of the opt-out test. Providing staff with clear, concise language to inform patients, particularly in situations where patient awareness is uncertain, may help staff communicate effectively without the need for lengthy consent discussions. It is important that training informs staff of situations (e.g., visual impairment or language barriers) in which the reminders may be helpful and appropriate, but that reminders are not necessary for every patient. This can help to normalise and streamline BBV screening in EDs [24].

Posters and other visual material informing patients about opt-out BBV testing should be on display in prominent positions throughout the ED and not lost among other posters [12]. This will not only ensure patients are informed but also reassure staff that patients are aware.

High workloads and competing priorities in EDs were a significant barrier to testing [24, 29]. However, there were specific systems and processes that facilitated the uptake of testing. Automation was found to support higher testing uptake, aligning with findings from previous studies [12, 15, 24, 32], although was not possible across all sites (two out of five sites did not have automation) IT systems could not facilitate automation, or additional resources were not available for set-up. By removing the need for staff to manually order tests, automation helped to streamline the process and reduced reliance on individual discretion. One study found that test uptake doubled when automation was introduced, compared to when nurses were required to order tests manually [33]. Automation also helped overcome barriers linked to stigma and risk profiling [15], contributing to the normalisation of BBV testing. By embedding testing into routine care, automation can help to reduce the influence of staff biases in deciding who should be tested [24].

Healthcare staff stigma towards people living with HIV remains an issue today [34]. This programme has the potential to shift perspective and reduce stigma by normalising BBV testing and increasing knowledge through the training provided in the programme.

Research has shown that education, including stigma reduction messaging, is a powerful tool for reducing HIV stigma which in turn would engage staff in the intervention [15, 24, 34].

Giving the intervention time to embed into practice and ensuring feedback loops and the flexibility to modify programme components was essential to sustaining the programme. Champions and key individuals appeared to play a vital role in maintaining staff motivation in the programme. These roles were at times ‘official’ roles but often relied on individual’s willingness and personal interest to drive the intervention. Key individuals facilitated effective communication between staff and created pathways for staff to provide feedback when issues arose, which facilitated adapting the intervention (e.g., a change in blood bottles). Data shows that although sites in the study were not consistently reaching 90% testing targets, staff nonetheless reported that they felt testing was embedded in practice [30]. This was in contrast to staff in one study who viewed the test as a ‘disruption’ [29]. Time may be a factor here, as interviews in that study were conducted early in the implementation phase, whereas some sites in the current study had been testing for over two years, highlighting the importance of allowing sufficient time for the intervention to embed [29].

Sites aiming to implement or scale up similar programmes should consider investing in automated test ordering, staff training, clear communication tools and dedicated individuals or champions who can maintain staff momentum and motivation (see Table 1). This can support the implementation of opt-out testing as an accepted part of emergency care that facilitates earlier diagnosis, reduces inequalities, and ultimately improves patient outcomes.

**Table 1.** Recommendations for Sites.

|  |  |
| --- | --- |
| **Implementation Planning** | 1. Foster early connections between hospital management, EDs, labs, HIV and Hepatitis services, IT teams, and clinical governance to streamline workflows and resolve issues efficiently.<br>2. Leverage national and regional NHSE networks by having a designated staff member attend meetings and sharing insights locally.<br>3. Engage staff early with regular updates, including testing figures and impactful case studies, to build understanding and sustain involvement.<br>4. Identify and support staff champions to drive momentum and promote programme adoption. |
| **Staff Training and Feedback** | 5. Provide accessible, practical, and knowledge-based training for all staff groups that includes content on the Programme's aims, local processes for test delivery, IT systems, care pathways for reactive results and up-to-date virus knowledge and stigma reduction.<br>6. Incorporate lived-experience stories in training to enhance staff understanding of the Programme's impact. |
| **Delivery of Testing** | 7. Share practical solutions to barriers from existing sites, such as modified vials and visual prompts, to improve test coverage [23].<br>8. Use simple scripts to reinforce the opt-out process and address staff concerns about patient communication. |
| **Reflection** | 9. Allow time for the programme to embed, enabling staff to adapt to new workflows.<br>10. Maintain ongoing communication about progress (including numbers of people detected, cost-effectiveness and contributions toward elimination plans) and provide opportunities for staff to share feedback to support long-term sustainability. |

Further research is needed to explore the patient experience of opt-out testing, including how those with different experiences and results (e.g. diagnosed, indeterminate, no result) respond to the intervention. This study found that outdated views on BBVs still existed amongst staff. Further in-depth exploration is needed, alongside the development of interventions aimed at increasing knowledge and reducing stigma in healthcare and the general public.

### Strengths and limitations

The sample in this study included a wide range of staff from HCAs to NHSE staff providing a varied and diverse view on implementation of the programme. The data was collected in 5 of the 32 sites, selected purposively to reflect geographic location, organisational practices, and opt-out testing processes, and only evaluated staff views of implementing the programme within ED. The use of NPT allowed for examination of issues with both the design of the intervention and its implementation. Interview data indicated that the NPT constructs of coherence and cognitive participation were well fulfilled for staff with positive consequences for implementation. Some findings may be site-specific and shaped by local characteristics or the length of time the programme had been running. It did not explore patients views and linkage to care, these will be explored in more detail in the wave 2 evaluation.

## Contributorship, in detailed form and the guarantor named

SA, TM, JH, RHT, LY, MH and SH conceived the study, SA and TM were responsible for collecting the data. SA, TM and JH were responsible for analysing the data. SA drafted the manuscript. The manuscript was reviewed and approved by SA, TM, JR, RHT, SH, MH, LY, RT, RH, and JH. The authors read and approved the final manuscript. Guarantor: JH

## Data Availability

All data produced in the present study are available upon reasonable request to the authors

## Acknowledgements, Funding, grant/award info

This research was funded by NHS England and the National Institute for Health Research (NIHR), with support from NIHR Health Protection Research Unit (HPRU) in Evaluation and Behavioural Science NIHR200877. The views expressed in this article are those of the authors and not necessarily those of NHS England, the NIHR or the Department of Health and Social Care.

## Ethical approval information, institution(s) and number(s)

This study obtained Oxford B Research Ethics Committee, HRA and HCRW approval (IRAS PROJECT ID 327922/REC Reference 23/SC/0357).

## Data sharing statement

The dataset is qualitative and includes numerous quotes from which participants are potentially identifiable. For this reason, the raw dataset will not be available. Additional quotes supporting each theme will be made available on request from Author

## Patient and Public Involvement

The study team participated in the programmes steering group, which included key individuals from the UK Health Security Agency (UKHSA), NHS England (NHSE), academia, community organisations involved with the care of people living with HIV and Hepatitis and people living with HIV and Hepatitis. These meetings provided a platform for the study team to share regular updates on study progress, discuss emerging challenges, and seek input on key aspects of the research. Study documents (including participant information, posters, and topic guides) were sent to all members of this group for comment and feedback.

## Abbreviations

BBV: Blood Borne Virus (HIV, Hepatitis B and Hepatitis C)
ED: Emergency Department
EPR: Electronic Patient Records
HCA: Health Care Assistant
HCV: Hepatitis C
HBV: Hepatitis B
HIV: Human Immunodeficiency Virus
NPT: Normalisation Process Theory
SOP: Standardised Operating Procedures

